# On the nutritional practices followed during containment, management and resolution of gastrointestinal fistulas. Results of a multicontinent, multinational, multicentric study

**DOI:** 10.1101/2022.11.02.22281854

**Authors:** Humberto Arenas Márquez, María Isabel Turcios Correia, Juan Francisco García, Roberto Anaya Prado, Arturo Vergara, Jorge Luis Garnica, Alejandra Cacho, Daniel Guerra, Miguel Mendoza Navarrete, Sergio Santana Porbén

## Abstract

**Introduction:** The multi-continent, multinational, multicenter “Fistula Day” Project has revealed a 14.7 % mortality rate in patients assisted for gastrointestinal fistulas (GIF) in hospitals of Latin America (LATAM) and Europe. GIF-associated mortality might be explained by the clinical-surgical condition of the GIF patient itself, the operational characteristics of the hospital, and surgical practices adopted locally in the containment, treatment and management of GIF. Given the hypermetabolic and cachectizing nature of GIF, it is then only natural to examine the nutritional practices adopted by medical teams in the care of GIF.

**Objective:** To examine the influence upon GIF outcomes of nutritional practices adopted in the hospitals participating in the “Fistula Day” Project.

**Study design:** Cohort study. During completion of the “Fistula Day” exercises 3 cross-sectional examinations were made: on admission in the study serie, and after 30 and 60 days after admission.

**Study serie:** One-hundred and seventy-seven patients (*Males*: 58.2 %; *Average age*: 51.0 ± 16.7 years; *Ages ≥ 60 years*: 36.2 %) assisted in 76 hospitals of Latin America (13 countries) and Europe (4).

**Methods:** The following nutritional practices adopted in the management of GIF were documented: assessment of the synthesis of hepatic secretory and acute phase proteins, patency of the oral route, administration of artificial nutrition, and use of glutamine as immunonutrient. Existence of a unit dedicated to clinical and hospital nutrition within the participating institution was also inquired. Impact of the documented nutritional practices upon survival of the patient, length of hospital stay, and spontaneous closure of the fistula was assessed.

**Results:** Rate of usage of nutritional practices was as follows (in descending order): *Determination of serum albumin*: 95.5 %; *Administration of artificial nutrition programs*: 80.8 %; *Existence of clinical and hospital nutrition unit*: 71.2 %; *Use of the oral route for sustaining the nutritional status of the patient*: 70.1 %; *Determination of C-reactive protein*: 36.1 %; and *Use of glutamine as immunonutrient*: 23.2 %; respectively. Rate of usage of nutritional practices was higher in patients with an enteroatmospheric fistula (EAF). In all the examined instances type of fistula determined GIF outcomes: enterocutaneous fistulas (ECF) were associated with higher likelihoods of survival and spontaneous closure, although at the expenses of prolonged hospital stay. Administration of the assessed nutritional practices only resulted in the prolongation of hospital stay. Existence of a unit dedicated to clinical nutrition was associated with reduced mortality, prolonged hospital stay and (numerically) lesser chance for spontaneous (non-surgical resolution) of GIF.

**Conclusions:** It is likely impact of the assessed nutritional practices to be mediated by the type of fistula, and that the benefit expected from a specified practice might be reduced (or even overruled) in patients assisted for EAF.

## INTRODUCTION

The “Fistula Day” Project has been completed in hospitals of Latin America (LATAM) and Europe with the main goal of documenting the practices currently followed in the containment and resolution of gastrointestinal fistulas (GIF).^1-3^ Impact upon GIF outcomes of the demographic characteristics of the patients,^1^ the operational characteristics of the participating hospital,^2^ and conducted surgical practices^3^ has been examined in successive publications.

Presence of an enteroatmospheric fistula (EAF) was associated with a higher mortality. On the other hand, hospital stay was dependent upon the type of surgery initially performed, location of fistula, and nutritional status of the patient (as estimated from calf circumference at admission in the study serie).^1^ None of the demographic characteristics of the GIF patient determined the spontaneous closure of the fistula.^1^

In a follow-up article, number of hospital beds emerged as the best predictor of survival of the GIF patient, prolongation of hospital stay, and a higher rate of spontaneous closure of the fistula.^2^ Existence of a unit specialized in intestinal failure and the number of patients assisted during one month might also influence (albeit marginally) upon evolution of GIF.

In the third article documenting the results of the “Fistula Day” Project the adoption of surgical practices for containment and resolution of GIF did not result in a higher GIF closure rate, although it is likely existence of a hospital unit specialized in the management of intestinal failure might bring about a higher rate of non-surgical closure of GIF.^3^

GIF are hypermetabolic and cachectizing events eventually leading to malnutrition, additional complications and death.^4-5^ Nutritional support should be a comprehensive part of the bundles of care provided to GIF patients. However, several reports have documented improper and untimely (if not delayed) nutritional practices in GIF patients thus resulting in reduced effectiveness and failure in achieving the established therapeutic goals.^6-7^

In addition to the surgical practices conducted in GIF patients, the design of the “Fistula Day” Project foresaw the documentation of the nutritional practices followed during the containment and resolution of GIF. The “Fistula Day” was extended to assess the impact of the documented nutritional practices upon management of GIF.

## MATERIAL AND METHOD

The design of the “Fistula Day” Project has been described in preceding publications.^1-3^ Briefly, hospitals eventually included in the project were invited to submit the demographical, clinical, sanitary, surgical and nutritional data of patients complicated with GIF between the months of May of 2019 and July of 2019 (both included) in three consecutive surveys.^1-3^ Participating hospitals were also surveyed on the presence within the institution’s organigram of a multidisciplinary unit dedicated to Clinical Nutrition (MDUCN).

The design of the “Fistula Day” Project foresaw the recording of nutritional practices adopted in the containment, treatment and eventual resolution of GIF, such as the determination of hepatic secretory (serum Albumin) and acute phase (C reactive Protein) proteins, patency of the oral route, administration of artificial nutrition, and use of glutamine as immunonutrient.

### Data processing and statistical-mathematical analysis of the results

Data submitted by the hospitals involved in the “Fistula Day” Project were entered into an on line application built upon *RedCap*®©^*^ (University of Vanderbilt, United States). The R program for statistical management and analysis (R Core Team 2018 version 3.5.0, United States) was used for debugging, preparing and processing data collected during the surveys of the “Fistula Day”. Data were reduced down to absolute | relative frequencies and percentages according with the type of the variable and the objective of the statistical analysis.

Condition of the patient (Alive/Deceased) upon discharge, hospital stay (Yes/No) and spontaneous closure of GIF (Yes/No) were assumed as the project outcomes at 30 and 60 days of the hospital’s admission in the “Fistula Day”. Nature and strength of the associations between the outcomes of the “Fistula Day” on one hand, and the identified nutritional practices, on the other; were examined by means of appropriate statistical *tests* in accordance with the type of the variable. Differences arising between cohorts of patients regarding the selected predictor were assessed by means of the log-rank test based in the chi-squared distribution.^8^ A level lower than 5% was used in all the instances to denote the finding as significant.

### Treatment of missing data

Data missing during follow-up of the patient were replaced with the observation recorded in the previous cross-sectional examination according with the LOCF (“Last Observation Carried Forward”) method.

### Intention-to-treat

Data gathered during the “Fistula Day” were analyzed according with the “Intention to treat” principle in order to keep fixed the size of the cohort.^9^

### Ethical considerations

The protocol followed by local surveyors during the “Fistula Day” was drafted in accordance with the “Good Clinical Practices” guidelines.^10^ Identity and rights of the surveyed patients were protected at all times.^11^ Patients (and by extension their caregivers) were informed about the purposes of the research, and the non-invasive nature of the procedures. Collected data were adequately preserved in order to ensure anonymity and confidentiality. Aggregated data were used solely for interpretation of the results and realization of statistical inferences. Informed consent was obtained from the GIF patient before inclusion in the cohort. Local conduction of the activities foreseen in the “Fistula Day” was authorized and supervised by the hospital Committees of Ethics after presentation, review and approval of the research protocols.

The researchers entrusted with the conduction of the “Fistula Day” Project presented the protocol “Current status of the postoperative fistula of digestive tract; multicentric, multinational study. DAY OF THE FISTULA” before the Ethics Committee of the San Javier Hospital (city of Guadalajara, State of Jalisco, México) for review and approval. A ruling was emitted on April 11^th^, 2018 by Dr. Eduardo Razón Gutiérrez, acting Director of the Ethics Committee, with the approval of the research protocol and the authorization for the conduction of the “Fistula Day” Project.

## RESULTS

### Main characteristics of the participating hospitals

Seventy-six hospitals from 17 countries participated in the “Fistula Day” activities.^1-3^ Thirteen of the participating countries were from LATAM. Sixty of the hospitals were Mexicans. Specialties hospitals prevailed (at least numerically). Most of the hospitals assisted between 1 – 2 GIF patients in a month-work. Participating hospitals distributed evenly regarding the number of beds. Most of the hospitals counted with an intensive care unit (ICU). In addition, three-quarters of the hospitals had a multidisciplinary unit dedicated to Clinical and hospital nutrition. On the contrary, a unit dedicated to the treatment of intestinal failure and/or postoperatory fistulas was only present in one-quarter of the hospitals. Expertise in GIF management of the acting physician was rated between “Expert” and “High” in one-third part of the hospitals.

### Main characteristics of the surveyed patients

One-hundred seventy-seven patients were surveyed during the “Fistula Day” exercises.^1-3^ Men prevailed over women accounting for 58.2 % of the size of the study serie. Average age was 51.0 ± 16.7 years. Subjects with ages ≥ 60 years represented 36.2 % of the studied cases. Fifty nine-point-six percent of the patients accumulated between 0 – 30 days of hospital stay at admission in the study serie. A diagnosis of cancer had been made in 27.7 % of the patients. Enterocutaneous fistula (ECF) was the prevailing type of fistula in the study serie. Almost 60 % of the GIF showed an output < 500 mL.day^-1^. Small bowel and colon were the dominant locations as origins of GIF. Half-plus-one of the GIF was diagnosed after the first 5 days of the primary surgery. In addition, 60.5 % of GIF originated after an emergency surgery.

### Main results of the “Fistula Day” Project

On conclusion of the present study the indicators of the evolution of GIF behaved as follows: *Mortality*: 14.7 %; *Prolonged hospitalization*: 46.3 %; and *Spontaneous closure of fistula*: 36.2 %.

### On the nutritional practices adopted in patients with gastrointestinal fistulas

Table 1 shows nutritional practices adopted by hospitals participating in the “Fistula Day”. Determination of serum Albumin had been made in 95.5 % of the instances. Determination of serum Albumin was independent from type of GIF: *EAF*: 96.8 % vs. *ECF*: 94.8 % (Δ = +2.0 %; p > 0.05; test for homogeneity based on the chi-square distribution). However, determination of C reactive Protein (CRP) was only made in one-third of GIF patients. Determination of CRP was also independent from type of fistula: *EAF*: 43.5 % vs. *ECF*: 32.1 % (Δ = +11.4 %; p > 0.05; test for homogeneity based on the chi-square distribution).

**Table 1.** Nutritional practices adopted during management of gastrointestinal fistulas. Results are presented as numbers and [within brackets] percentages of patients assessed in each practice. Assessed nutritional practices are also shown for each type of fistula. Legend: AN: Artificial nutrition.

| *Adopted nutritional practice* | *Enteroatmospheric*<br>*fistulas* | *Enterocutaneous*<br>*fistulas* | *All the fistulas* |
| --- | --- | --- | --- |
| Size | 62 [35.0] | 115 [65.0] | 177 [100.0] |
| Determination of Serum albumin | 60 [96.8] | 109 [94.8] | 169 [95.5] |
| Determination of C reactive Protein | 27 [43.5] | 37 [32.2] | 64 [36.1] |
| Suspension of the oral route | 19 [30.6] | 34 [29.6] | 53 [29.9] |
| Administration of AN programs | 59 [95.2] | 84 [73.0] | 143 [80.8] |
| Use of glutamine as immunonutrient | 19 [30.6] | 22 [19.1] | 41 [23.2] |
| Existence of a Clinical and hospital<br>nutrition unit | 49 [79.0] | 77 [66.9] | 126 [71.2] |

Size of the study serie: 177.
Source: Records of the study.

Oral route was suspended in less than one-third of GIF patients at the moment of admission in the project. Suspension of oral route was independent from type of GIF: *EAF*: 30.6 % vs. *ECF*: 29.6 % (Δ = +1.0 %; p > 0.05; test for homogeneity based on the chi-square distribution).

Artificial nutrition (AN) programs had been administered in 80.8 % of GIF patients included in the “Fistula Day”. Rate of administration of NA programs was higher in EAF patients: *EAF*: 95.2 % vs. *ECF*: 73.0 % (Δ = +22.2 %; χ^2^ = 12.697; p < 0.05; test for homogeneity based on the chi.-square distribution).

Glutamine was used as immunonutrient in nearly a fifth of GIF patients. Use of glutamine was independent from type of fistula: *EAF*: 30.6 % vs. *ECF*: 19.1 % (Δ = +11.5 %; p > 0.05; test for homogeneity based on the chi-square distribution).

A clinical and hospital nutrition unit (CHNU) existed in 71.2 % of the instances. Existence of the CHNU was independent from type of fistula: *EAF*: 79.0 % vs. *ECF*: 66.9 % (Δ = +12.1 %; p > 0.05; test for homogeneity based on the chi-square distribution).

### Impact of nutritional practices upon survival of the patient

Determination of serum CRP was associated with condition of GIF patient upon discharge from the “Fistula Day”: survival rate was lower among patients in whom CPR was determined: *Survival rate*: CRP determined: 78.1 % *vs*. CRP not determined: 89.4 % (Δ = -11.3 %; p < 0.05; test for homogeneity based on the chi-square distribution). Condition of the GIF patient upon discharge from the “Fistula Day” was also associated with the existence of a CHNU: survival rate was (again) lower among patients assisted in hospitals in which a CHNU existed: *Survival rate*: Unit existing: 81.7 % vs. Unit not existing: 94.1 % (Δ = -12.3 %; p < 0.05; test for homogeneity based on the chi-square distribution).

In contrast with these findings, the remaining nutritional practices did not influence upon survival of GIF patients: *Survival rate*: Determination of serum Albumin: Serum albumin determined: 85.2 % *vs*. Serum albumin not determined: 87.5 % (Δ = -2.3 %; p > 0.05; test for homogeneity based on the chi-square distribution); Suspension of oral route: Oral route suspended: 86.8 % *vs*. Oral route not suspended: 84.7 % (Δ = +2.1 %; p > 0.05; test for homogeneity based on the chi-square distribution); Administration of AN programs: AN programs administered: 83.9 % *vs*. AN programs not administered: 91.2 % (Δ = -7.3 %; p > 0.05; test for homogeneity based on the chi-square distribution); and Use of Glutamine as immunonutrient: Glutamine used: 85.4 % vs. Glutamine not used: 85.3 % (Δ = +0.1 %; p > 0.05; test for homogeneity based on the chi-square distribution).

Impact of nutritional practices upon survival of GIF patients was adjusted for type of fistula by means of logistic regression techniques. Exception made for the determination of serum Albumin (OR_Fistula type_ = 1.597; 95 % CI: 0.729 – 3.500; p > 0.05 vs. OR_Determination Albumin_ = 1.663; CI 95 %: 0.857 – 3.225; p > 0.05), survival of GIF patient was dictated primarily by the type of fistula: likelihood of survival was higher for patients having an ECF: *Determination of CRP*: OR_Fistula type_ = 2.969; 95 % CI: 1.685 – 5.230; p < 0.05 vs. OR_Determination CRP_ = 0.958; 95 % CI: 0.525 – 1.747; p > 0.05; *Suspension of oral route*: OR_Fistula type_= 2.141; 95 % CI: 1.126 – 4.069; p < 0.05 vs. OR_Oral route_ = 1.340; 95 % CI: 0.736 – 2.438; p > 0.05; *Administration of AN programs*: OR_Fistula type_ = 2.139; 95 % CI: 1.150 – 3.977; p < 0.05 vs. OR_AN Administration_ = 1.318; 95 % CI: 0.766 – 2.367; p > 0.05; *Use of Glutamine as immunonutrient*: OR_Fistula type_ = 2.034; 95 % CI: 1.146 – 3.613; p < 0.05 vs. OR_Glutamine use_ = 1.609; 95 % CI: 0.765 – 3.388; p > 0.05; and *Existence of CHNU*: OR_Fistula type_ = 2.964; 95 % CI: 1.552 – 5.659; p < 0.05 vs. OR_CHNU_ = 1.000; 95 % CI: 0.569 – 1.755; p > 0.05; respectively.

Figure 1 shows the impact of nutritional practices upon survival of GIF patients after disaggregating the study serie in the corresponding cohorts. Of the assessed nutritional practices, only the existence and operation of the CHNU influenced upon survival rate of each of the cohorts subjected to comparison: survival rate was always lower among those GIF patients treated in hospitals having a CHNU: *30 days-survival*: Existence of CHNU: 86.5 % vs. No Existence of CHNU: 96.1 %; *60 days-survival*: Existence of CHNU: 81.7 % (Δ = +4.8 %) vs. No Existence of CHNU: 94.1 % (Δ = +2.0 %; log-rank test χ^2^ = 4.076; p < 0.05). Determination of CRP also influenced (albeit marginally) survival rates of the cohorts: *30 days-survival*: CPR determined: 84.4 % vs. CPR not determined: 92.0 %; *60 days-survival*: CPR determined: 78.1 % (Δ = +6.3 %) vs. CPR not determined: 89.4 % (Δ = +2.6 %; log-rank test χ^2^ = 3.698; p = 0.0545).

**Figure 1.**
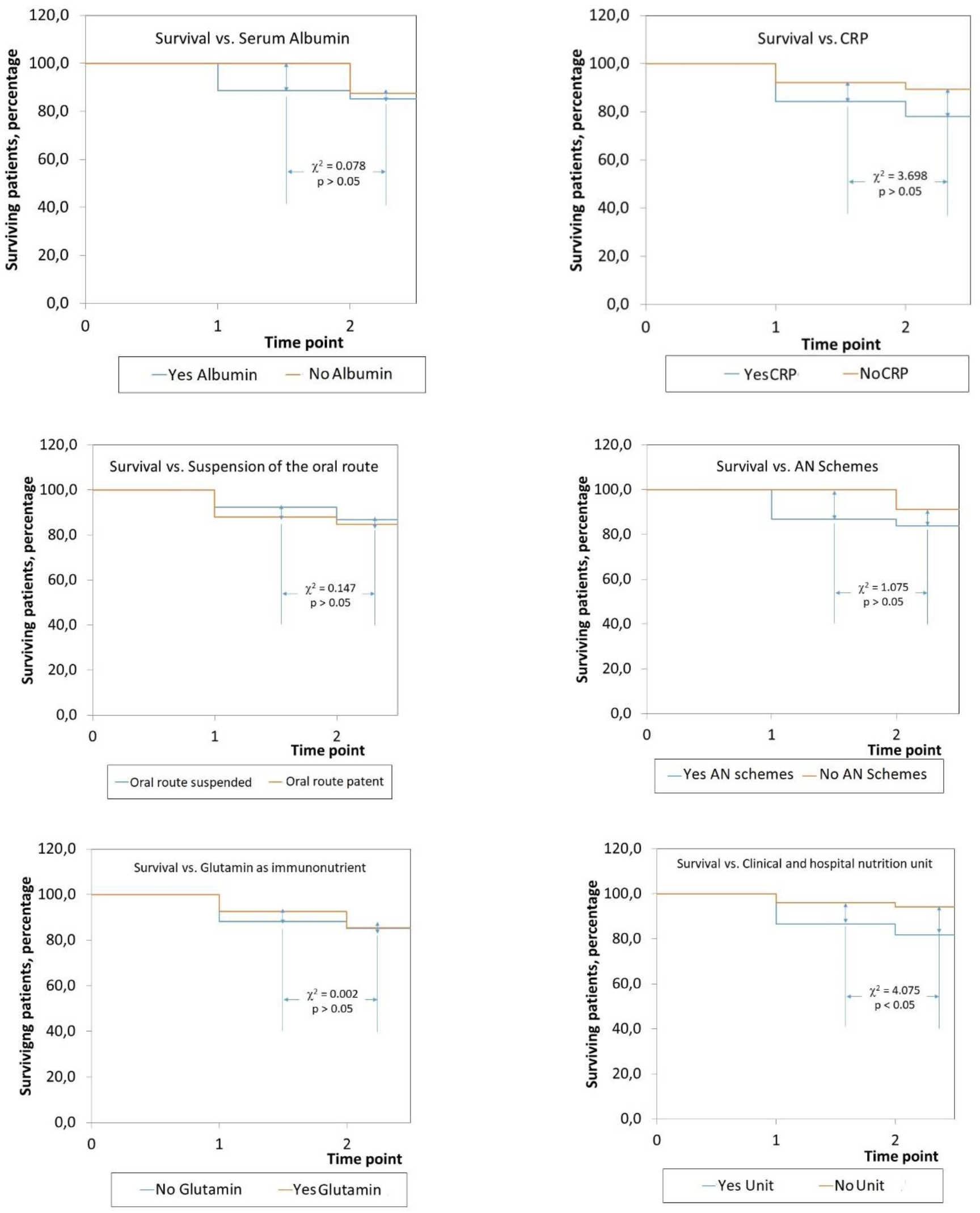
Impact of assessed nutritional practices upon survival of patients with gastrointestinal fistulas. The study serie was dessagregated into the corresponding cohorts. Legend: CRP: C reactive Protein. AN: Artificial nutrition. Size of the study serie: 177. Source: Records of the study.

### Impact of nutritional practices upon length of hospital stay

Impact of assessed nutritional practices upon hospital length of stay was heterogeneous. Administration of AN programs, use of Glutamine as immunonutrient, and existence of a unit dedicated to clinical and hospital nutrition were associated with prolongation of hospital stay: Prolonged hospital stay: *Administration of AN programs*: AN programs administered: 53.1 % *vs*. AN programs not administered: 23.5 % (Δ = +29.6 %; p < 0.05; test for homogeneity based on the chi-square distribution); *Use of Glutamine as immunonutrient*: Glutamine used: 70.7 % *vs*. Glutamine not used: 40.4 % (Δ = +30.3 %; p < 0.05; test for homogeneity based on the chi-square distribution); *Existence of a unit dedicated to clinical and hospital nutrition*: Existence of the unit: 53.9 % *vs*. No existence of the unit: 31.3 % (Δ = +22.6 %; p < 0.05; test for homogeneity based on the chi-square distribution).

Hospital stay was prolonged in patients in whom serum albumin was not determined: *Serum albumin determined*: 45.6 % vs. *Serum albumin not determined*: 87.5 % (Δ = -41.9 %; p < 0.05; test for homogeneity based on the chi-square distribution). Interestingly, hospital stay was prolonged in those patients in whom oral route was not suspended: *Oral route not suspended*: 54.8 % vs. *Oral route suspended*: 30.2 % (Δ = +24.6 %; p < 0.05; test for homogeneity based on the chi-square distribution).

In contrast with the previously presented findings, prolongation of hospital stay was independent from determination of CRP: CRP determined: 46.8 % *vs*. CRP not determined: 47.8% (Δ = -1.0 %; p > 0.05).

Impact of nutritional practices upon prolongation of hospital stay of the GIF patients was further adjusted for the type of fistula. Both EAF and adopted nutritional practice explained a higher likelihood of prolongation of hospital stay in three of the instances: *Administration of AN programs*: OR_AN Administration_: 1.547 (95 % CI: 1.003 – 2.383; p < 0.05) *vs*. OR_Fistula type_: 0.601 (95% CI: 0.377 – 0.959; p < 0.05); *Use of glutamine as immunonutrient*: OR_Glutamine use_: 1.980 (95 % CI: 1.143 – 3.431; p < 0.05) *vs*. OR_Fistula type_: 0.580 (95 % CI: 0.390 – 0.889; p < 0.05); and *Existence of CHNU*: OR_CHNU_: 1.560 (95 % CI: 1.001 – 2.431; p < 0.05) *vs*. OR_Fistula type_: 0.609 (95% CI: 0.385 – 0.963; p < 0.05).

There was no impact of determination of serum protein upon hospital stay: *Determination of serum Albumin*: OR_Determination Albumin_ = 0.724 (95 % CI: 0.431 – 1.215; p > 0.05) vs. OR_Fistula type_ = 1.361 (95 % CI: 0.754 – 2.456; p > 0.05); *Determination of CRP*: OR_Determination CRP_ = 1.014 (95% CI: 0.635 – 1.621; p > 0.05) vs. OR_Fistula type_ = 0.923 (95 % CI: 0.622 – 1.368; p > 0.05).

GIF patients subjected to suspension of the oral route showed a higher likelihood of shorter hospital stays: a finding opposing the type of fistula: OR_Oral route_= 0.573 (95 % CI: 0.336 – 0.979; p < 0.05) vs. OR_Fistula type_= 1.389 (95 % CI: 0.912 – 2.119; p = 0.059).

Figure 2 shows the impact of the adopted nutritional practices upon hospital stay of GIF patients after constructing the corresponding cohorts of cases. Of the surveyed nutritional practices, only use of Glutamine as immunonutrient influenced upon the behavior of the cohorts: hospitalization rates were higher in GIF patients in whom Glutamine was used: *30 days-hospitalization*: Glutamine used: 46.3 % *vs*. Glutamine not used: 32.4 %; 6*0 days-hospitalization*: Glutamine used: 24.4 % (Δ = +21.9 %) *vs*. Glutamine not used: 8.1 % (Δ = +24.3 %; *log-rank test* = 4.179; p < 0.05).

**Figure 2.**
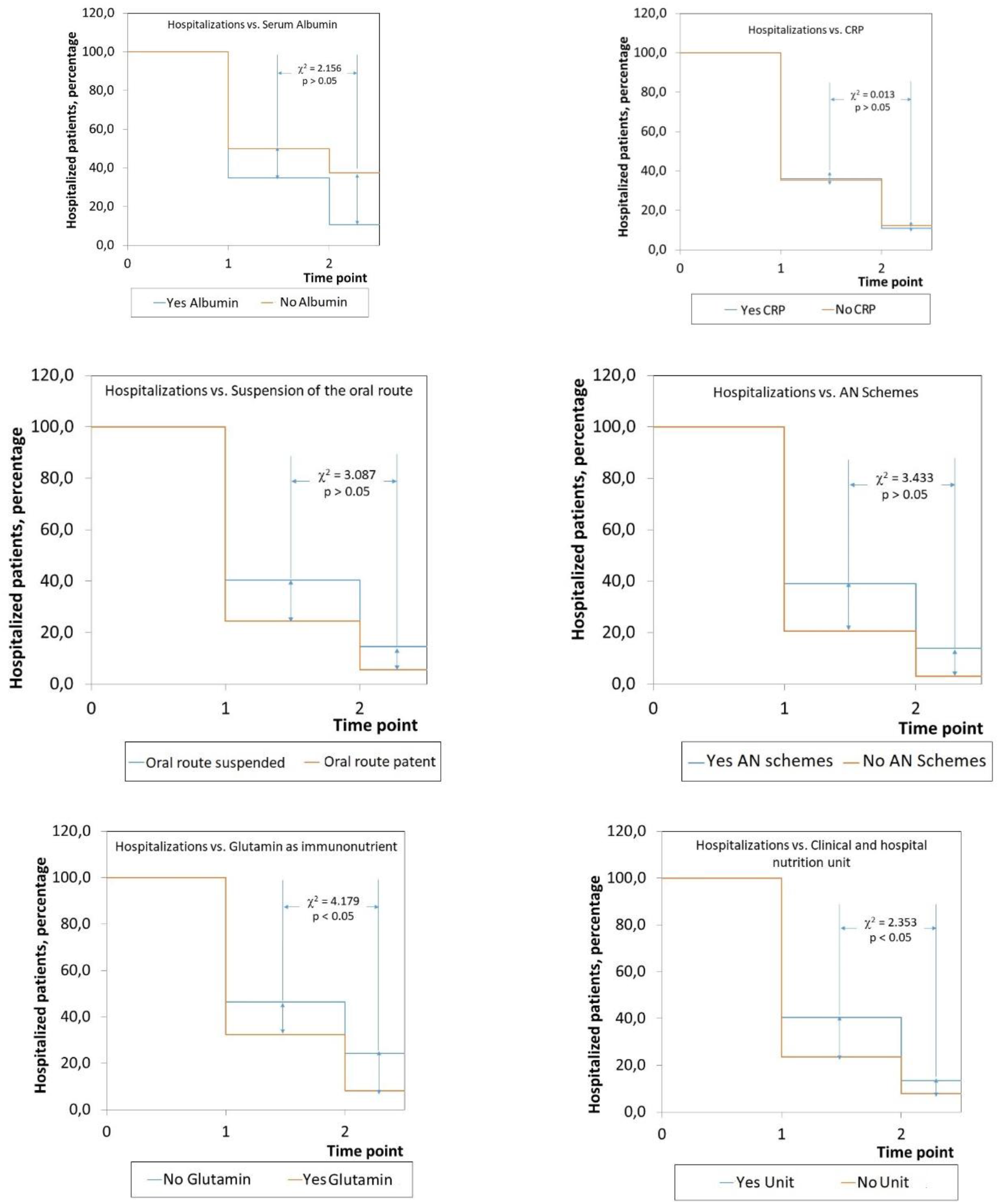
Impact of assessed nutritional practices upon hospital length of stay in patients with gastrointestinal fistulas. The study serie was dessagregated into the corresponding cohorts. Legend: CRP: C reactive Protein. AN: Artificial nutrition. Size of the study serie: 177. Source: Records of the study.

### Impact of nutritional practices upon spontaneous closure of the fistula

Successful closure (that is: without further refistulization) of the fistula is the ultimate goal of nutritional practices adopted in the GIF patient. Twenty-five events of refistulization were counted during the window of observation of the study.

Of all the adopted nutritional practices spontaneous closure of fistula was dependent only upon the status of the oral route: events of refistulization were more frequent in those GIF patients with a suspended oral route: *Suspended oral route*: 93.1 % vs. *Oral route not suspended*: 80.8 % (Δ = +12.3 %; p < 0.05; homogeneity test based on the chi-square distribution).

Impact of the assessed nutritional practices upon spontaneous closure of GIF was adjusted for type of fistula. In most of the instances spontaneous closure of fistula was only dependent upon type of fistula, with ECF showing the highest likelihood (data not shown). Exception is to be made of the determination of CRP: spontaneous closure of GIF was most likely in those GIF patients in whom CRP was determined: OR_Determination CRP_: 0.498 (95 % CI: 0.247 – 1.004; p < 0.05) *vs*. OR_Fistula type_: 0.587 (95 % CI: 0.332 – 1.036; p < 0.05).

In this regard, it is worth to be noticed existence of a CHNU influenced (albeit marginally) upon spontaneous closure of fistula: OR_CHNU_: 0.642 (95 % CI: 0.361 – 1.143; p = 0.062) *vs*. OR_Fistula type_: 0.532 (95 % CI: 0.289 – 0.979; p < 0.05).

Figure 3 presents the impact of the adopted nutritional practices upon the spontaneous closure of GIF when the study serie was disaggregated in the corresponding cohorts. None of the nutritional practices influenced upon the behavior of the resulting cohorts (data not shown).

**Figure 3.**
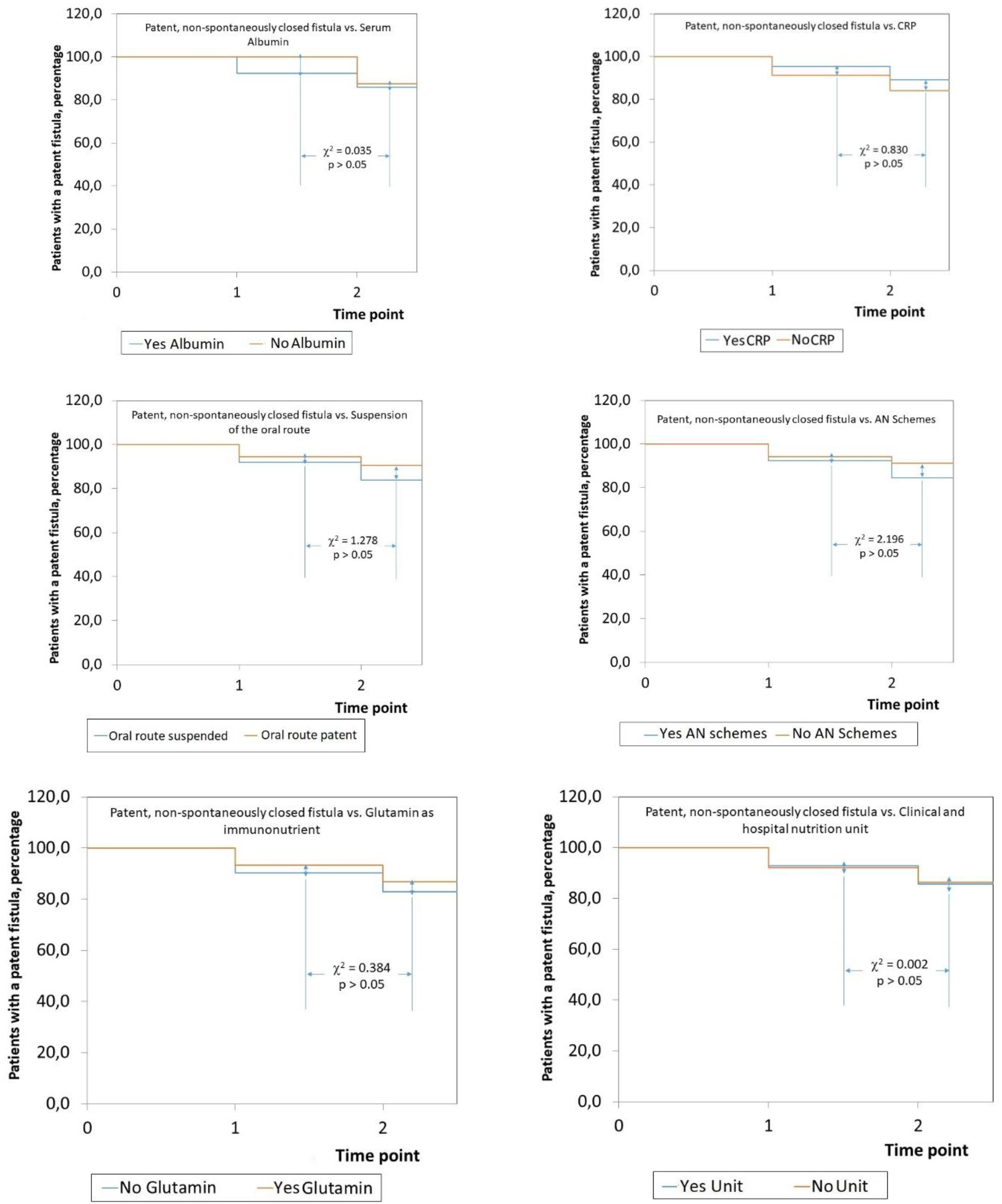
Impact of assessed nutritional practices upon spontaneous closure of gastrointestinal fistulas. The study serie was dessagregated into the corresponding cohorts. Legend: CRP: C reactive Protein. AN: Artificial nutrition. Size of the study serie: 177. Source: Records of the study.

### Characteristics of the administered Artificial Nutrition programs

Finally, Table 2 shows the characteristics of the AN programs administered to GIF patients. Central parenteral nutrition prevailed as the most reported AN program in the study serie. Parenteral nutrition was compounded as an individualized order. Mixed AN programs (parenteral + enteral nutrition) were administered to a fifth of the GIF patients. An enteral access was placed in one out of eight GIF patients. A surgical ostomy was the prevalent enteral access. AN programs were used to provided 26 – 30 kcal.kg^-1^.day^-1^ of energy (32.2 % of the study serie) and 1.1 – 1.5 g.kg^-1^.day^-1^ of proteins (44.6 % of the study serie) in most of the GIF patients.

**Table 2.** Characteristics of the nutritional artificial programs administered to patients with gastrointestinal fistulas.

| Characteristic | Findings |
| --- | --- |
| ***Mode of artificial nutrition*** |  |
| • Enteral nutrition | 27 [15.3] |
| • Central Parenteral Nutrition | 74 [41.8] |
| • Peripheral Parenteral Nutrition | 6 [ 3.4] |
| • Mixed: Parenteral + Enteral | 36 [20.3] |
| • Not administered / Not declared | 34 [19.2] |
| ***Mode of parenteral nutrition*** |  |
| • Standardized | 34 [19.2] |
| • Individualized | 81 [45.8] |
| • Not declared | 62 [35.0] |
| ***Enteral access*** |  |
| • Not placed | 147 [83.1] |
| • Placed | 30 [16.9] |
| ***Type of acces placed <sup>¶</sup>*** |  |
| • Nasogastric | 9 [30.0] |
| • Nasojejunal | 8 [26.7] |
| • Surgically placed | 13 [43.3] |
| ***Energy provided, kcal.kg<sup>-1</sup>.day<sup>-1</sup>*** |  |
| • < 15 | 1 [ 0.6] |
| • 15 – 20 | 25 [14.1] |
| • 21 – 25 | 45 [25.4] |
| • 26 – 30 | 57 [32.2] |
| • > 30 | 3 [ 1.7] |
| • No declared | 46 [26.0] |
| ***Proteins provided, g.kg<sup>-1</sup>.day<sup>-1</sup>*** |  |
| • 0.5 – 1.0 | 17 [ 9.6] |
| • 1.1 – 1.5 | 79 [44.6] |
| • 1.6 – 2.0 | 1 [ 0.6] |
| • > 2.0 | 48 [27.1] |
| • No declared | 32 [18.1] |

<sup>¶</sup> Regarding those 30 patients in whom an enteral access was placed.
Size of the study serie: 177.
Source: Records of the study.

## DISCUSSION

The present work has described the nutritional practices administered to GIF patients surveyed during the “Fistula Day” exercises. As such, this work complements and extends the results presented in previous publications regarding the demographic, clinical and sanitary characteristics of the surveyed GIF patients, as well as the hospital practices conducted on them as part of GIF management.^1-3^

On average, nutritional practices were administered to merely half of GIF patients. In addition, nutritional practices were used mostly in EAF patients. It was not the purpose of the present work to examine the causes for the observed results. Determination of both hepatic secretory and acute-phase proteins is customary for assessing the nutritional status of the GIF patients as well as persistence of pro-inflammatory states.^6^ Given the hypercatabolic and cachectizing nature of GIF, all patients with this condition should be assisted by a unit specialized in clinical and hospital nutrition and also entrusted with the design, implementation and conduction of the required AN programs.^14^ In this regard, it is to be noticed AN programs were administered to a higher proportion of EAF patients, leading to speculate there are differences depending on the type of fistula in the perceptions the medical care teams have about the impact of GIF upon the nutritional status of the patient and the need for nutritional care and support as part of their medical and surgical management.

It is still striking one third of GIF patients had the oral route suspended on the occasion of the “Fistula Day” activities. Suspension of the oral route is no longer recommended for the management of GIF, being a practice resulting in aggravation | perpetuation of clinical and nutritional derangements brought about by fistulas themselves.^6-7^ In addition, suspension of oral route does not translate to a reduction in the fistula output.^6-7^

Immunonutrition has been promoted as a specialized nutritional intervention in several surgical scenarios, with glutamine being an important metabolic substrate of the enterocyte.^12^ It would be then natural to use nutritional formulas containing glutamine to promote a better outcome in the management of GIF. In this regard, Martinez *et al*. (2019) supplemented ECF patients awaiting definitive surgery with glutamine (10 g.day^-1^) + arginine (4.5 g.day^-1^) during 7 days.^12^ Supplemented patients showed a lower rate of refistulization as well as no post-surgical infections.^12^ Mean concentrations of serum IL-6 and CRP were also lower in the supplemented patients.^12^ However, immuno-enhancing nutritional formulas might be not only expensive but also not available to all GIF patients, and their prevalent use in EAF patients might signal to a heroic action conducted by the medical care team in order to achieve resolution of the fistula when other practices have failed.

Most of the nutritional practices assessed in this work did not influence upon survival of GIF patients. On the other hand, it was striking determination of CRP and existence of a CHNU were both associated with a lower survival rate of GIF patients. In addition, survival was only dependent upon type of fistula, with ECF exhibiting the highest likelihood of survival. It was beyond the purpose of the “Fistula Day” to provide explanations for these associations given the observational nature of the project. However, lower survival rate of GIF patients subjected to these practices might be due, in part, to the (delayed) timing of their implementation, when they can hardly benefit from them.

Impact of nutritional practices upon length of hospital stay was mixed. Certain practices determined a shorter hospital stay, while others resulted in prolongation of the stay of GIF patients. Administration of AN programs and use of glutamine led to prolonged stay, implying time has to be allocated for the nutritional practice adopted in the GIF patient to be implemented during hospitalization and its effectiveness assessed. Hospital stay might be further prolonged if nutritional practices are adopted in EAF patients, in particular when EAF have been associated in several studies with a poor prognosis.^13^

The present work found suspension of the oral route distinguished for a higher likelihood of shortened hospital stay opposing type of fistula. It was not the purpose of this work to provide explanations for such a finding. Only one third of GIF patients were subjected to this practice. In contrast, prolonged hospitalizations concentrated among those patients with preserved oral route. Paucity of data might then have been a reason for this finding. On the other hand, suspension of the oral route might have been a temporary intervention taken in several patients irrespective of the type of the fistula discharged early otherwise.

It would be expected the assessed nutritional practices might lead to a higher rate of spontaneous closure of GIF. However, none of them resulted in a better GIF outcome. In this regard, it is to be noticed refistulization was more frequent in GIF patients in whom oral route was suspended, providing additional evidence about the futility of this practice. In addition, the present work showed the higher likelihood of spontaneous closure of ECF independently from the nutritional practice adopted.

Existence of a CHNU was marginally associated with a higher likelihood of GIF spontaneous closure: a relevant finding given the heterogeneity of the study serie. CHNU are implemented with the purpose of providing both general and specialized nutritional care to hospitalized patients by a multidisciplinary team and in accordance with evidence-based recommendations.^14^ However, impact of the CHNU upon spontaneous closure of GIF might be mediated by a prolongation of hospital stay, raising concerns among administrators and managers about the costs of GIF management.

## CONCLUSIONS

At present, assessed nutritional practices do not impact upon GIF outcomes. Nutritional practices were not of universal application, and possibly administered preferably to EAF patients in an untimely fashion and as heroic actions. It is to be noticed suspension of the oral route was associated with a higher rate of refistulization. Administration of nutritional practices to GIF patients might imply allocation of additional time and resources, thus increasing the costs of hospitalization and casting doubts about their effectiveness on health directives and managers.

## Data Availability

The researchers declare their willingness and disposition to share the research data with those interested.

## Footnotes

* Available at: http://www.redcap.org.

